# Chronic adaptations following eccentric cycling training at different cadences

**DOI:** 10.64898/2026.07.07.26356024

**Authors:** Adrien Mater, Alain Martin, Davy Laroche, Romuald Lepers

## Abstract

Pedalling cadence during an acute eccentric cycling exercise altered physiological and perceptual responses. We examined the influence of cycling cadence on neuromuscular adaptation induced by a 6-week eccentric cycling training period. Eighteen participants performed training (eighteen sessions) at a cadence of 30 or 60 rpm over six weeks. Power output was the same between the two groups. Perceived effort and heart rate were recorded at each training session. Muscle pain and fatigue were reported the day after each session. Maximal voluntary contractions torque, as well as concentric and eccentric cycling efficiency, were assessed before and after training. Additionally, the loss of maximal voluntary isometric torque was assessed after the first and last training sessions. Heart rate and perceived effort increased in the second week of training and then plateaued, with no difference between groups. Muscle pain and fatigue remained low throughout the training, with no difference between groups. Isometric (+28%) and eccentric (+13%) maximal voluntary torque of knee extensor muscles increased regardless of training cadence. Concentric maximal voluntary torque increased for the group pedalling at 60 rpm only (+21%). Cycling efficiency was improved in eccentric mode only (+43%), with no difference between the two training groups. Finally, the voluntary isometric torque loss induced by the first and last sessions were similar. While six weeks of eccentric cycling training improved neuromuscular and functional capacities, cadence had no observable effect. This finding suggest that patients could choose their preferred cadence to obtain better adherence to the rehabilitation program without altering the adaptations.

## Introduction

Cycling is probably the most prescribed endurance exercise in rehabilitation programs. Several clinical trials have shown that it can improve patients’ quality of life and functional abilities, such as muscle strength (25). It is possible to adapt the workload of training sessions by varying the duration and intensity of the exercise and changing the contraction mode. In this context, some ergometers allow the isolation of the eccentric contraction during a pedaling movement (40). Briefly, the lower limb muscles engage in eccentric contractions while resisting the backward motion of the cranks produced by an ergometer. This exercise modality reduces energy demand and perceived exertion compared to concentric cycling performed at a similar power output (10). Despite that eccentric contractions can induced muscle damage, it has been shown that eccentric pedaling can be gradually implemented to limit delayed onset muscle soreness (13, 33) and dyspnea symptoms during exercise in chronic obstructive pulmonary disease patients (32).

Furthermore, eccentric cycling can be performed at higher power outputs than concentric cycling without inducing a high cardiorespiratory demand (10, 20, 21). Indeed, when an eccentric versus concentric cycling training period is performed at the same heart rate, power output increased faster for eccentric and reached almost four times the power out developed in concentric cycling at the end of 8 weeks of training (22). Moreover, similar results have been reported when concentric and eccentric cycling are compared and defined at the same perception of effort (upon 6-20 Borg’s Scale)(6, 20) or for similar oxygen consumption (30). In these cases, cycling is performed at a higher power output in the eccentric mode than in the concentric mode. The greater mechanical stress imposed during eccentric cycling training is an interesting stimulus for locomotor muscle strengthening and hypertrophy. Indeed, eccentric cycling allows for greater gains in isometric, isokinetic (concentric and eccentric) strength of the knee extensor muscles and functional performance (6-minute walking test) compared to those observed after conventional concentric cycling training (7, 20, 22, 23, 27, 34).

Studies on cycling defined the exercise based on intensity through the intensity – power output. However, modulating cadence in – conventional or eccentric - cycling also leads to different cardiovascular demands, perceptions of effort, and muscle activation (2, 29, 30) for a given power output. In eccentric cycling, frequently used in rehabilitation programs, the objective is to physical capacities while limiting the difficulty of exercises (i.e., perceived exertion and oxygen consumption). In this sense, the research on cadence has sought to achieve a compromise between these two parameters. There is a cadence around 60 rpm, called the optimal cadence, which minimizes perceived effort, oxygen consumption, and muscle activation (28). Shifting upward or downward from the optimal cadence increased acute physiological and perceptual responses. Thus, as eccentric cycling allows the production of a greater power output than conventional cycling, reducing the cadence would allow an increase in the force applied to the pedals.

The present study aimed to compare neuromuscular and functional adaptations following a training period of 6 weeks of eccentric cycling performed at two different cadences but similar power outputs. Moreover, it aimed to describe neuromuscular adaptations induced by eccentric cycling by dissociating central from peripheral adaptations. We assumed that training at a cadence of 30 rpm would lead to greater neuromuscular adaptations but would be perceived as harder than training at 60 rpm.

## Method

### Participants

Twenty volunteers (7 females and 13 males) participated in this study. Two participants (1 male and 1 female) did not complete all training sessions due to an injury occurring in their free time during the training period. Thus, data from eighteen participants were finally analyzed (table 1). Participants were free from injuries in their lower limbs during the last 3 months. Most participants were novices to eccentric (ECC) cycling and only practiced concentric cycling for daily commutes. Only four of them participated in a short ECC cycling experiment (3 sessions with various cadences) one year before the present one. Based on the criteria presented by two studies (11, 12), participants were defined as recreationally trained.

**Table 1:** Anthropometric characteristics, Peak power output (PPO) and peak oxygen consumption (VO_2_peak) of participants in the two groups. Each group were composed with three females and six males.

|  | *ECC<sub>30</sub>* (*n* = 9) |  | *ECC<sub>60</sub>* (*n* = 9) |  |
| --- | --- | --- | --- | --- |
| *Age (years)* | 23 ± 3 |  | 22 ± 3 |  |
| *Height (cm)* | 177.6 ± 9.3 |  | 173.8 ± 9.3 |  |
|  | *Pre* | *Post* | *Pre* | *Post* |
| *Mass (kg)* | 63.6 | 63.9 | 66.1 | 66.1 |
| *PPO (W.kg<sup>-1</sup>)* | 3.8 ± 0.6 | 3.9 ± 0.6 | 3.9 ± 0.9 | 3.8 ± 0.9 |
| *VO<sub>2peak</sub>*<br>*(ml.min<sup>-1</sup>.kg<sup>-1</sup>)* | 49.5 ± 9.9 | 50.1 ± 10.0 | 50.7 ± 5.9 | 49.9 ± 8.2 |

**Table 2:** Internal load parameters throughout training weeks. * indicates different from week 1, one sign means different at p < 0.05, three signs mean different at p < 0.001.

| Variable | Condition | Effect | Week 1 | Week 2 | Week 3 | Week 4 | Week 5 | Week 6 |
| --- | --- | --- | --- | --- | --- | --- | --- | --- |
| Heart rate<br>(% peak) | ECC<sub>30</sub> | TIME*** | 58.7 ± 9.1 | 57.9 ± 6.2 | 61.1 ± 7.4 | 62.0 ± 8.0 | 66.7 ± 9.6 | 64.0 ± 9.4 |
|  | ECC<sub>60</sub> |  | 56.2 ± 9.7 | 53.3 ± 8.3 | 55.3 ± 5.5 | 58.9 ± 5.5 | 63.1 ± 7.0 | 63.2 ± 7.6 |
| Perception<br>of effort (a.u.) | ECC<sub>30</sub> | TIME*** | 22 ± 7 | 26 ± 12 | 30 ± 13 | 33 ± 10 | 42 ± 19 | 36 ± 16 |
|  | ECC<sub>60</sub> |  | 30 ± 10 | 26 ± 8 | 28 ± 9 | 34 ± 9 | 40 ± 12 | 31 ± 12 |
| DOMS post 24h<br>(a.u.) | ECC<sub>30</sub> | No effect | 17 ± 8 | 11 ± 11 | 15 ± 9 | 16 ± 8 | 22 ± 11 | 15 ± 9 |
|  | ECC<sub>60</sub> |  | 22 ± 12 | 16 ± 12 | 16 ± 11 | 17 ± 12 | 14 ± 12 | 13 ± 11 |
| Fatigue post<br>24h (a.u.) | ECC<sub>30</sub> | No effect | 16 ± 7 | 18 ± 8 | 22 ± 9 | 23 ± 11 | 22 ± 11 | 20 ± 16 |
|  | ECC<sub>60</sub> |  | 22 ± 11 | 22 ± 10 | 24 ± 9 | 23 ± 7 | 22 ± 10 | 26 ± 11 |

The study was approved by the French ethics committee (ClinicalTrials.gov Identifier: NCT04886115), conducted in accordance with the declaration of Helsinki (2008) and all participants signed a written informed consent.

### Experimental design

Participants were enrolled in the study depending on their availability for about 9 weeks including a 6-week training period (Figure 1). They were allocated to one of two groups using a pseudo-randomized method considering their concentric cycling peak power output (PPO) and isometric maximal voluntary contraction (MVC_ISO_) torque. Participants in both groups had to maintain their physical activity and dietary habits as far as possible compared to the two months before the experimentation. Participants’ group characteristics are presented in Table 1.

**Figure 1:** Overview of the experimental protocol. Shapes with a grey background illustrate the cycling training period performed by both experimental groups at 30 or 60 rpm (ECC_30_ and ECC_60_, respectively). FAM = familiarization, performed 10 to 13 days before the start of training, includes an incremental concentric cycling test (PPO test) and a familiarization for eccentric cycling (EEC) and neuromuscular function assessment (NM function); PRE = pre-test, includes the assessment of muscular architecture (ultrasound), neuromuscular function and concentric cycling efficiency (CONC CE); the first and last training sessions, includes a neuromuscular function test and eccentric cycling efficiency realized before and after the eccentric cycling exercise; INT = during training test, includes assessment of neuromuscular function; POST = post-test, performed 7 to 10 days after the end of training includes all assessment performed in previous sessions.

All participants take part in different testing sessions before (i.e., a familiarisation and a pre-test) and after (post-tests) the training period. During the familiarization session, the volunteers performed a continuous incremental cycling test (starting at 50 W, increasing by 1 W every 3 s) on the concentric ergometer until exhaustion, allowing the determination of their PPO and peak oxygen consumption (VO2_PEAK_). Then, participants were introduced to the neuromuscular testing procedure, perceptual scales, and eccentric cycling during 20-min at 60%PPO at a cadence corresponding to their allocated group (30 or 60 rpm). Furthermore, all measurements were explained to the participants. This session was performed 10 to 13 days before the first session of training allowing participants to recover from the eccentric cycling bout. The pre-test session (PRE) was located three days before the start of the training period. This session started with the assessment of muscular architecture at rest. Then, participants warmed up at 40% PPO for 10 min on the concentric ergometer. Lastly, pre-training neuromuscular function and concentric cycling efficiency were performed at 60%PPO and the freely chosen cadence was assessed. An intermediate test (INT) was performed on the same day of the 10^th^ session before eccentric cycling to assess neuromuscular function in the middle of the training period. The post-test (POST) was spread over two sessions on distinct days. The first post-session was composed of measures of muscular architectural and incremental concentric cycling tests. In contrast, the second one included neuromuscular testing procedure and concentric cycling efficiency performed at 60%PPO (PRE training) and freely chosen cadence. The POST measurements were placed 7 to 10 days after the end of training to avoid residual fatigue.

### Training period

Eccentric cycling consists of resisting against a motorized semi-recumbent cycle-ergometer adapted to impose active muscle lengthening of knee extensor muscles with a backward rotation of the cranks (Cyclus 2, Cyclus GmbH, Leipzig, Germany). Participants were asked to match distinct cadences in *isopower* mode according to their eccentric cycling group. ECC_30_ and ECC_60_ refer to the group in which participants had to maintain a cadence of 30 and 60 revolutions per minute (rpm), respectively. Eighteen sessions were distributed over the six weeks at the rate of three sessions per week. Training load increased in time from 20 to 30-min of cycling at 60%PPO in the first two weeks. Then, while duration remained constant, power output increased by 15%PPO per week until 105%PPO and remained similar the last two weeks.

### Internal load during training

Perception of effort (i.e., difficulty breathing and driving legs to pedal)(27) and delayed onset muscle soreness (DOMS) of the knee extensor muscles (i.e., how much it globally hurt in their leg muscles), and perception of fatigue using Cr100 scale of Borg (6). During training, the perception of effort was reported at the last minute of each training session. Moreover, DOMS and perceived fatigue were reported by participants 24 h after each training session.

### First and last training sessions

The first (1^st^) and last (18^th^) training sessions allowed us to evaluate oxygen consumption during exercise and exercise-induced neuromuscular alterations. The design of both sessions is based on that used in the study by a previous study of our team (29). Briefly, neuromuscular tests were realized before and after eccentric cycling performed at 30 or 60 rpm, depending on the groups. According to the general design of the training, participants had to perform a 20-min and 30-min exercise at 60 and 105%PPO for the 1^st^ and the 18^th^ sessions, respectively. The cardiorespiratory function was assessed throughout the exercise duration.

### Cardiorespiratory function

The wearable metabolic system (K5, Cosmed, France) was calibrated before each use according to company guidelines. The mask was associated with the face of the participants just before the start and removed directly after the end of the cycling exercise. The oxygen uptake (VO_2_, ml.min^-1^.kg^-1^), respiratory frequency (RF, bpm), and minute ventilation (VE, l.min^-1^) were recorded during the incremental concentric tests and during both 1^st^ and 18^th^ sessions. The heart rate (HR, bpm) was measured using a chest strap heart rate monitor (HR Module, Garmin, United States of America) paired with the metabolic system.

### Neuromuscular function

Participants were seated on an isokinetic dynamometer (System pro 4, Biodex Medical System, New York) with a hip flexed at 90° and the knee joint axis in line with the rotation axis of the dynamometer, the leg strapped to the lever arm 2 cm above the malleoli, and the torso strapped against the seat. Percutaneous electrical stimulations were delivered on the right right femoral nerve using a high-voltage constant-current stimulator (model DS7, Digitimer, Hertfordshire, UK) via a ball probe cathode (0.5 cm diameter) manually pressed on the femoral nerve using a stylus. A self-adhesive anode (10 x 5 cm) was placed in the gluteal fossa. Stimulation intensity was progressively increased until both M-wave amplitudes of vastus lateralis and rectus femoris muscles and the torque responses plateaued. We then used 150% of this intensity (0.5ms, 400V) to assess maximal M-wave (M_MAX_) and associated muscle twitch properties.

Participants performed maximal voluntary isometric knee extensor contractions (MVC_ISO_) with their right leg. Then, they performed isokinetic maximal voluntary concentric (MVC_CONC_) and eccentric (MVC_ECC_) knee extensor contractions over a 90° range of motion between 95 and 5° of amplitude (with 0° corresponding to full leg extension) and at a velocity of 60°.s^-1^. At least two MVCs of each contraction mode were performed (more if the difference between the peak torque of the two contractions was greater than 5%). The trial with the greatest peak torque was analyzed. Neuromuscular function was completed using percutaneous peripheral electrical stimulations. A double stimulation at 100 Hz was triggered manually during the torque plateau of each MVC_ISO_. Two more double stimulations (100 Hz-Dt100 and 10 Hz- Dt10, respectively) and one simple stimulation (Tw) were delivered at rest after the MVC_ISO_ (31). Also, participants performed a Thorstensson Test (39). Briefly, in the same configuration as dynamic maximal voluntary contractions, participants had to repeat fifty MVC_CONC_ at 180°.s^-1^. Participants were instructed to give an all-out effort during each concentric action and to relax their leg during flexion back to the starting position at same angular velocity.

Lastly, the testing procedure for jump performance imitates the recommendation set to assess jump height (3). Participants performed three jumps of both squat jump (SJ) and countermovement jump (CMJ) separated with 1-min recovery. The trial with the greatest height was analyzed. Participants had to keep their hands on their hips, starting from a static standing position, leg straight for CMJ and leg fold for SJ. Participants were instructed to jump as high as possible. Jump performance was assessed using *My jump* app with Iphone 7 (Apple Inc., USA). One participant did not perform the POST training jump test because he twisted his ankle two days before.

### Muscular architecture

Muscle ultrasound imaging was performed on the vastus lateralis and rectus femoris muscles of the right leg using a B-mode ultrasonography system (Z.one SmartCart, Zonare medical system INC). Muscular imaging was assessed longitudinally using a 6-cm linear array ultrasound probe at the midpoint of the muscle length according to the two medial and discal musculotendinous junctions of each muscle. Participants lay in supine position with their legs fully extended. Both medial and distal myotendinous junctions were found to determine muscle length. Thus, three screenshots were taken in the middle of each muscle. Three different measures of muscle thickness and pennation angle of three fascicles were directly reported during assessment. Then fascicles length was estimated according to pennation angle and muscle thickness. Patellar tendon stiffness was assessed during maximal isometric knee extension contractions performed at a knee joint angle of 90°, using a linear 3-s ramp from rest to maximal voluntary contraction. Tendon elongation was simultaneously measured using real-time ultrasonography. The ultrasound probe was kept fixed to the skin at the level of the distal musculotendinous junction of the vastus lateralis muscle throughout the entire duration of the contraction. Tendon stiffness was calculated as the slope of the tendon force–elongation relationship over the force interval between 50 and 100% of MVC.

### Data analysis

Workload (J) was calculated as the product of session duration and power output of each participant. Maximal torque amplitude was analyzed for Dt100, Dt10, single twitch and MVC before and after cycling exercises. The rate of torque development (RTD, N.s^-1^) was calculated by dividing the torque value by contraction time, while the half-relaxation rate (HRR, N.s^-1^) was measured from the peak torque plateau to the point it fell to half its peak value. Both variables were the ratio between torque change and the corresponding time. Strojnik & Komi formula was used to obtain the voluntary activation level (VAL), where the torque at the moment of stimulation is compared with the superimposed twitch torque (38). MMAX duration (peak to peak), amplitude, and area for RF and VL muscles were investigated. RMS EMG during MVC was measured during 500 ms around the peak torque and divided by MMAX area (EMG RMSMAX/MMAX) to estimate the magnitude of voluntary neural drive. For each participant, the slope of torque decrease throughout the Thorstensson test was determined using the director coefficient of function. Moreover, the voluntary maximal muscle torque performed during the Thorstensson test was reported as MVC_CONC_ performed at 180°.s^-1^. Jump ratio was calculated between gains in CMJ and SJ heights and considered as the capacity to use leg stiffness after the training period. While muscle thickness was reported for both vastus lateralis and rectus femoris muscles, pennation angle and fibers length were reported for vastus lateralis only. The cycling efficiency (CE) of the 1st and the 18th session was calculated for eccentric as well as the concentric cycling efficiency assessed before and after the training period. CE included the power output (PO, in W), the time of exercise (t), the oxygen consumption (VO2, in L.min-1), and the energetic equivalent associated to respiratory ratio (O2 equiv, in kJ.L-1) associated to respiratory ratio (14).

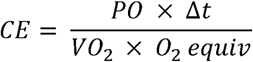

### Statistics

Because of the low number of participants in each group, data were not filtered to remove outlier. Statistical analysis was performed with the software JAPS (version 0.16.1; JASP TEAM, Amsterdam, Netherlands). A Shapiro-Wilk test determined whether the data followed a normal distribution. Thus, two-way repeated measures ANOVA served to assess the effect of GROUP (ECC_30_ and ECC_60_), TIME (PRE, INT and POST) and GROUP x TIME interaction for maximal voluntary contractions, PPO and VO_2PEAK_. Cycling efficiency and jump height changes were compared between training groups only (ECC_30_, ECC_60_) using two-way repeated measures ANOVA (CADENCE x TIME). Another two-way repeated measures ANOVA (CADENCE x TIME) was used to follow heart rate, perception of effort, muscle pain, and perceived fatigue collected during training sessions (six weeks) for the training groups (ECC_30_, ECC_60_). Greenhouse-Geisser’s correction was applied when the data did not conform to the assumption of sphericity. Then, the Holm post-hoc test followed significant ANOVA results, and Cohen’s d completed the P value of followed-up analyses. All data are expressed as mean ± SD.

## Results

### Participants characteristics

All the anthropometric characteristics of the participants did not differ between groups (table 1). Peak power output (W/kg) and peak oxygen consumption were not affected by TIME or GROUP (all p > 0.28, all η_p_^2^ < 0.07).

### Internal load parameters during training sessions

Training sessions are presented in Table 1. Heart rate and perception of effort increased over TIME (all p < 0,001, all η_p_^2^ > 0,397) without CADENCE effect (all p > 0.16, all η_p_^2^ < 0.10). Perception of muscle pain and fatigue reported 24h after each session remained stable throughout the training weeks without any impact of the CADENCE (all p > 0.073, all η_p_^2^ < 0.123).

### Functional parameters

MVC of knee extensor muscles performed in isometric (Figure 2, panel A), and eccentric (Figure 2, panel B) concentric (Figure 2, panel C) performed at 60°.s^-1^ were conducted three times during experiment (PRE, INT, POST) while concentric contractions performed at 180°.s^-1^ (Table 3) were performed twice (PRE, POST).

**Figure 2:** Evolution of maximal voluntary contractions following training period. MVC torque was assessed in isometric (panel A), eccentric (panel B), and concentric (panel C) at 60°.s-1 before (PRE), during the third week (INT), and after six weeks (POST) of the training period. Data were present with bar averaged for both groups when repeated measure ANOVA did not show CADENCE x TIME interaction. * The above bars indicate the difference from PRE. # above bars indicate differences from INT. One sign means different at p < 0.05, two signs mean different at p < 0.01, and three signs mean different at p < 0.001.

**Table 3:**
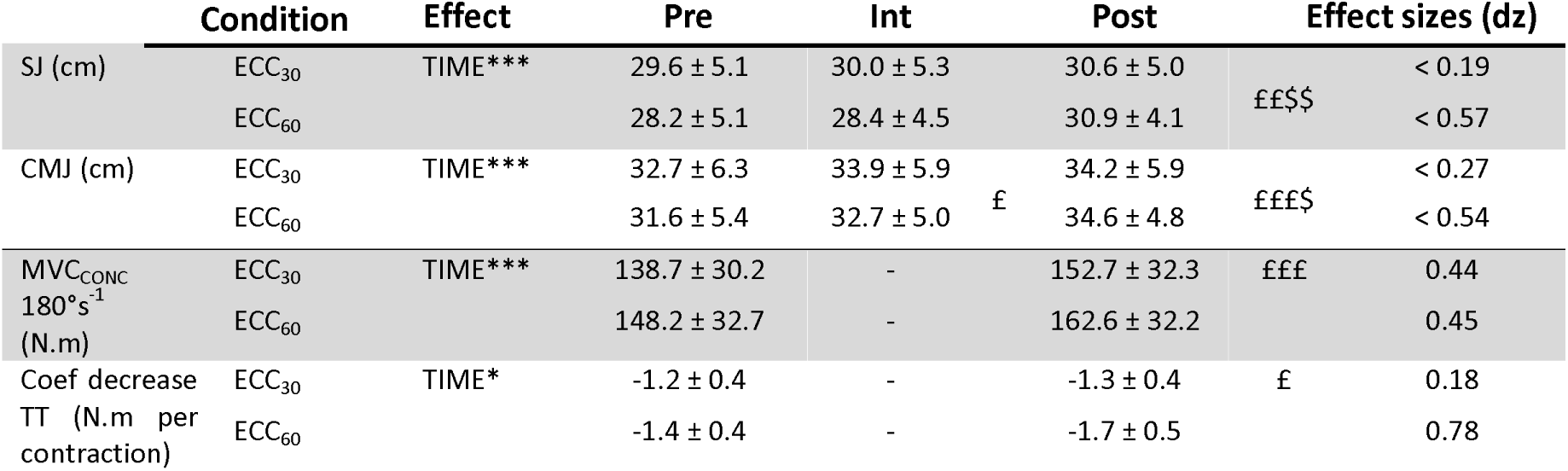
Height of squat jump (SJ), countermovement jump (CMJ), and Thorstensson test (MVC_CONC_ 180°s and coefficient decrease during Thorstensson test (TT)) parameters for both experimental groups performed before (PRE), during (INT) and after (POST) the training period. *means a main effect in the repeated measure ANOVA, £ means different from Pre value, $ means different from Int value. One sign means p < 0.05, two signs mean p < 0.01, three signs mean p < 0.001.

MVC_ISO_ increased with TIME without difference between CADENCE (p < 0.01, η_p_^2^ = 0.60). Post hoc analyses revealed significant increases between all-time points (all p < 0.03, all d > 0.45). MVC_CONC_ performed at 60°.s^-1^ showed a TIME x GROUP interaction effect (p < 0,05, η_p_^2^ = 0,18). Post-hoc analysis showed that voluntary concentric torque was greater at INT and POST compared to PRE for ECC_60_ (p < 0,05, d > 0.66) but remained stable in ECC_30_ (all p > 0.99, all d < 0,30). MVC_ECC_ showed a main effect of TIME (p < 0,05, η_p_^2^ = 0,08). Post-hoc analysis showed that voluntary isometric torque increased at all-time intervals regardless of CADENCE (all p < 0.05, all d > 0.31). MVC_CONC_ performed at 180°.s^-1^ showed only a main effect of TIME (p < 0,05, η_p_^2^ = 0,08).

SJ and CMJ height (Table 3) increased with TIME (all p < 0.001, all η_p_^2^ > 0.37) with no effect of CADENCE (all p > 0.08, all η_p_^2^ < 0.16).

### Neuromuscular function

Neuromuscular function evaluated through peripheral stimulations applied during and after MVC_ISO_ was presented in Table 4. None of the parameters were affected by CADENCE (all p >0.26, all η_p_^2^ > 0.08).

**Table 4:** Changes in neuromuscular function parameters before (PRE); during (INT) and after (POST) cycling exercise. Dt100: doublet 100Hz, Dt10: doublet 10Hz, Tw: single simulation, RTD: rate of torque development, Mmax: maximal M-wave responses to single stimulation, $ Mean increase for both conditions compared to pre-value.

| Variable | Condition | Effect | PRE | INT | POST | Effect size |
| --- | --- | --- | --- | --- | --- | --- |
| VAL (%) | ECC30 | TIME* | 88.5 ± 9.0 | 92.3 ± 6.0 | 91.8 ± 5.9 | < 0.58 |
|  | ECC60 |  | 89.6 ± 8.4 | 92.0 ± 5.1 | 94.4 ± 3.4 | < 0.72 |
| EMG<sub>MAX</sub> RMS/M<sub>MAX</sub><br>Vastus lateralis | ECC30 | No effect | 0.067 ± 0.032 | 0.046 ± 0.023 | 0.051 ± 0.020 | < 0.40 |
|  | ECC60 |  | 0.061 ± 0.021 | 0.066 ± 0.017 | 0.090 ± 0.109 | < 0.57 |
| EMG<sub>MAX</sub> RMS/M<sub>MAX</sub><br>Rectus femoris | ECC30 | TIME* | 0.074 ± 0.038 | 0.081 ± 0.031 | 0.113 ± 0.056 | \$ < 0.93 |
|  | ECC60 |  | 0.093 ± 0.049 | 0.105 ± 0.034 | 0.116 ± 0.038 | < 0.54 |
| Dt100 amplitude<br>(N.m) | ECC30 | TIME* | 73.4 ± 19.8 | 79.5 ± 17.5 | 78.1 ± 21.7 | \$ < 0.34 |
|  | ECC60 |  | 73.1 ± 15.4 | 75.0 ± 14.6 | 81.5 ± 17.6 | < 0.47 |
| Dt100 RTD<br>(N.s<sup>-1</sup>) | ECC30 | No effect | 800 ± 333 | 702 ± 200 | 756 ± 213 | < 0.40 |
|  | ECC60 |  | 696 ± 196 | 736 ± 222 | 756 ± 277 | < 0.26 |
| Dt10 amplitude<br>(N.m) | ECC30 | No effect | 66.2 ± 20.5 | 72.1 ± 22.0 | 65.0 ± 23.3 | < 0.37 |
|  | ECC60 |  | 65.4 ± 11.4 | 71.6 ± 16.1 | 74.8 ± 18.9 | < 0.49 |
| Dt10 RTD<br>(N.s<sup>-1</sup>) | ECC30 | No effect | 411 ± 124 | 428 ± 134 | 425 ± 127 | < 0.15 |
|  | ECC60 |  | 399 ± 53 | 442 ± 104 | 455 ± 103 | < 0.59 |
| Tw<br>(N.m) | ECC30 | No effect | 44.3 ± 13.1 | 48.8 ± 12.4 | 41.8 ± 1 | < 0.56 |
|  | ECC60 |  | 41.4 ± 7.5 | 45.5 ± 12.3 | 44.0 ± 13.5 | < 0.32 |
| M<sub>MAX</sub> duration<br>Vastus lateralis (ms) | ECC30 | INTERACTION* | 9.4 ± 2.9 | 10.1 ± 2.8 | 7.1 ± 1.7 | < 1.02 |
|  | ECC60 |  | 8.4 ± 3.3 | 8.2 ± 2.8 | 10.7 ± 3.5 | < 0.89 |
| M<sub>MAX</sub> amplitude<br>Vastus lateralis (mV) | ECC30 | No effect | 4.9 ± 2.9 | 6.0 ± 3.3 | 6.5 ± 2.7 | < 0.49 |
|  | ECC60 |  | 6.4 ± 3.3 | 6.6 ± 3.2 | 7.2 ± 3.4 | < 0.27 |
| M<sub>MAX</sub> area<br>Vastus lateralis (mV.ms) | ECC30 | No effect | 32 ± 18 | 39 ± 21 | 40 ± 14 | < 0.47 |
|  | ECC60 |  | 37 ± 21 | 41 ± 17 | 43 ± 19 | < 0.34 |
| M<sub>MAX</sub> duration<br>Rectus femoris (ms) | ECC30 | No effect | 11.8 ± 3.2 | 11.1 ± 2.4 | 9.5 ± 1.9 | < 0.96 |
|  | ECC60 |  | 10.7 ± 1.9 | 11.4 ± 3.1 | 10.7 ± 1.6 | < 0.32 |
| M<sub>MAX</sub> amplitude<br>Rectus femoris (mV) | ECC30 | No effect | 3.2 ± 1.0 | 3.0 ± 1.1 | 2.5 ± 0.9 | < 0.48 |
|  | ECC60 |  | 3.6 ± 1.4 | 3.7 ± 1.9 | 3.3 ± 1.9 | < 0.31 |
| M<sub>MAX</sub> area Rectus femoris (mV.ms) | ECC30 | No effect | 19 ± 7 | 17 ± 9 | 15 ± 5 | < 0.51 |
|  | ECC60 |  | 20 ± 7 | 22 ± 9 | 21 ± 13 | < 0.22 |

Voluntary activation level increased with TIME (p < 0.05, η_p_^2^ = 0.18). EMG RMS/M of vastus lateralis did not show any effect of TIME or GROUP (all p > 0.39, all η_p_^2^ < 0.05). EMG_MAX_ RMS/M_MAX_ of rectus femoris showed a main effect of TIME (p < 0.05, η_p_^2^ = 0.21). Post hoc showed that EMG_MAX_ RMS/M_MAX_ of rectus femoris was greater in POST than PRE values (p > 0.05, d = 0.73).

Torque evoked by Dt100 increased with TIME (p < 0.05, η_p_^2^ = 0.20) whereas torque induced by the 10-Hz doublet (Dt10) and single twitch remained unchanged (all p > 0.99, all d < 0,30). The rate of torque development of Dt100 and Dt10 did not show any effect of TIME (all p > 0.16, all η_p_^2^ < 0.11).

### Muscle architecture and tendinous stiffness

Muscle adaptations are presented in Table 5. Vastus lateralis and rectus femoris muscle thickness increased after the training period (all p < 0.05, all η_p_^2^ > 0.32) but were unaffected by CADENCE (all p > 0.40, η_p_^2^ < 0.04). Vastus lateralis pennation angle and muscle fascicles showed no effect of TIME or CADENCE (all p > 0.09, η_p_^2^ < 0.17). Patellar tendon stiffness increased from 358.6 ± 150.1 to 390.0 ± 176.6 Nm.cm^-1^ following training (p < 0.05, η_p_^2^ = 0.27) with not effect of CADENCE (p > 0.23, d < 0,09).

**Table 5:**
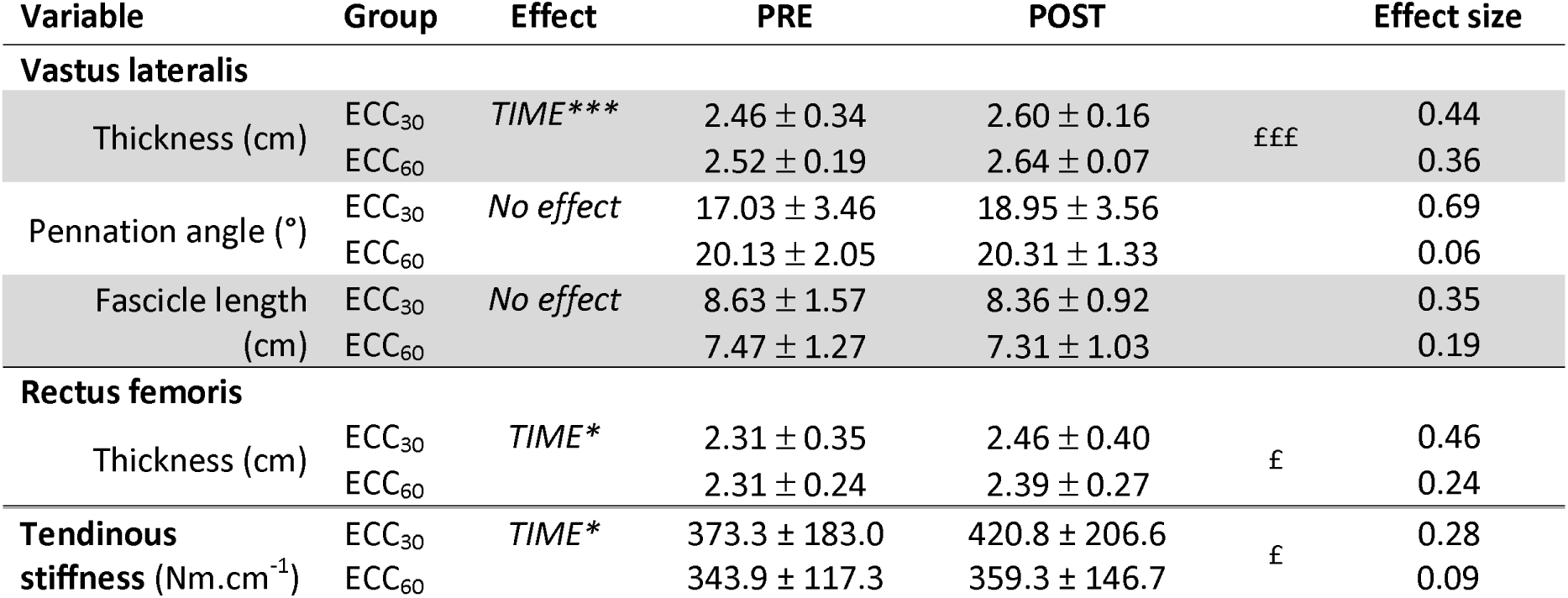
Change in architecture of the vastus lateralis and rectus femoris muscles and patellar tendon stiffness. * means a main effect in the repeated measure ANOVA, £ means different from Pre value, one sign means different at p < 0.05, three signs mean different at p < 0.001.

| Variable | Group | Effect | PRE | POST |  | Effect size |
| --- | --- | --- | --- | --- | --- | --- |
| Vastus lateralis |  |  |  |  |  |  |
| Thickness (cm) | ECC<sub>30</sub> | TIME*** | 2.46 ± 0.34 | 2.60 ± 0.16 | £££ | 0.44 |
|  | ECC<sub>60</sub> |  | 2.52 ± 0.19 | 2.64 ± 0.07 |  | 0.36 |
| Pennation angle (°) | ECC<sub>30</sub> | No effect | 17.03 ± 3.46 | 18.95 ± 3.56 |  | 0.69 |
|  | ECC<sub>60</sub> |  | 20.13 ± 2.05 | 20.31 ± 1.33 |  | 0.06 |
| Fascicle length (cm) | ECC<sub>30</sub> | No effect | 8.63 ± 1.57 | 8.36 ± 0.92 |  | 0.35 |
|  | ECC<sub>60</sub> |  | 7.47 ± 1.27 | 7.31 ± 1.03 |  | 0.19 |
| Rectus femoris |  |  |  |  |  |  |
| Thickness (cm) | ECC<sub>30</sub> | TIME* | 2.31 ± 0.35 | 2.46 ± 0.40 | £ | 0.46 |
|  | ECC<sub>60</sub> |  | 2.31 ± 0.24 | 2.39 ± 0.27 |  | 0.24 |
| Tendinous stiffness (Nm.cm<sup>-1</sup>) | ECC<sub>30</sub> | TIME* | 373.3 ± 183.0 | 420.8 ± 206.6 | £ | 0.28 |
|  | ECC<sub>60</sub> |  | 343.9 ± 117.3 | 359.3 ± 146.7 |  | 0.09 |

### Thorstensson test

Strength loss during the Thorstensson test increased from 1.29 to 1.50 N·m per contraction between PRE and POST for both groups (p < 0,05, η_p_^2^ = 0,35) with no effect of CADENCE (p = 0,13, η_p_^2^ = 0,17).

### First and last training sessions

Power output increased from 60 to 105% PPO between the first and the last session. Oxygen consumption increased with TIME (p < 0,01, η_p_^2^ = 0,53) but was not affected by CADENCE (p = 0,55, η_p_^2^ = 0,03) (figure 3, panel A). MVC torque loss after the 1^st^ and the 18^th^ sessions did not show any effect of TIME or CADENCE (all p > 0.2, all η_p_^2^ < 0.10) (figure 3, panel B).

**Figure 3:** Oxygen consumption (panel A, n = 14) and MVC torque loss (panel A, n = 18) induced by the first (1st) and the last session (18th) performed at 60 and 105%PPO, respectively. Box plots represent the minimum, the 25th percentile, the median, the mean (+), the 75th percentile, and the maximum.

### Cycling efficiency

Cycling efficiency showed a main interaction effect of TIME x MODALITY (p < 0.001, η_p_^2^ = 0.76). Concentric cycling efficiency was lower than eccentric (p < 0.001, η_p_^2^ = 0.97). Concentric cycling efficiency was stable between the first and last training sessions (p = 0.89, d = 0.05). In contrast, eccentric cycling efficiency increased between the first and last sessions (p < 0.001, d = 2.81) without any effect of CADENCE (p > 0.05, η_p_^2^ = 0.07; figure 4).

**Figure 4:** Concentric and eccentric cycling efficiencies performed on semi-recumbent cycle-ergometer before/after and at the 1st/18th sessions, respectively. Both ECC30 and ECC60 groups were average in a single value because no CADENCE x TIME interaction effect was found. ***indicates a difference in eccentric cycling efficiency during the 18th compared to the 1st session at p < 0.001. Box plots represent the minimum, the 25th percentile, the median, the mean (+), the 75th percentile, and the maximum.

## Discussion

This is the first study comparing the effects of eccentric cycling training performed at two different cadences on neuromuscular function. We hypothesized that because acute physiological responses were increased at 30 compared to 60 rpm, functional capacity would be improved to a greater extent after training at 30 rpm. Contrary to this hypothesis, most of the variables assessed were not influenced by cadence. The only exception was knee extensor MVC_CONC_ torque at 60°·s□¹, which increased exclusively after training at 60 rpm. In addition, internal training load indicators (i.e., heart rate and perceived exertion) did not differ between groups throughout the intervention.

Previous studies reported higher internal load responses (oxygen consumption, heart rate, and perceived exertion) when eccentric cycling was performed at low cadences (30 rpm) compared with intermediate cadences (60 rpm) (19, 28, 29). In contrast, internal load was similar between cadences in the present study (Table 2). This discrepancy may be explained by differences in participant familiarization: in earlier studies, a single familiarization session may not have been sufficient to fully accustom participants to eccentric pedaling, whereas repeated exposure during training may have attenuated cadence-related differences. Furthermore, the present comparison was conducted between individuals assigned to two groups exercising within the same intensity domain (moderate intensity)(8), rather than using an intra-individual design, which may have limited the detection of subtle differences.

The relatively low cardiorespiratory demand of eccentric cycling in physically active individuals (32–38% VO□max during the first and last training sessions, respectively; Figure 3) likely explains the absence of improvements in PPO or VO□max. Consistent with previous findings, eccentric pedaling appears insufficient to elicit maximal cardiorespiratory adaptations in healthy populations, whereas improvements in cardiovascular function have been reported primarily in clinical populations (4). Nevertheless, eccentric cycling efficiency was higher than concentric efficiency before training, as originally described by the team of Abbott in 1952, and increased further only when cycling was performed in the eccentric mode (Figure 4)(1). This eccentric-specific improvement in efficiency may reflect a progressive optimization of pedaling technique for a novel motor task and/or an increased contribution of passive musculotendinous components, which could reduce metabolic cost during eccentric contractions. Although patellar tendon stiffness increased following training (Table 5), it was not assessed during eccentric contractions, and therefore this interpretation should be considered with caution. Additionally, hypertrophy of the knee extensors (increased VL and RF thickness) may have increased total capillary number without changes in mitochondrial or capillary density—adaptations more typically associated with endurance training (18, 22) — thereby contributing to improved efficiency.

Despite the progressive increase in workload, internal load (heart rate and perceived exertion), delayed onset muscle soreness, and perceived fatigue remained low and stable throughout the training period (Table 2). These findings suggest that workload progression was sufficiently gradual to avoid excessive muscle damage (23, 24, 33). Future studies could complement subjective assessments with objective markers such as blood creatine kinase and additional functional tests. Importantly, in a rehabilitation context, minimizing excessive fatigue is essential to preserve daily functional capacity. In this regard, despite a substantial increase in total workload between the first (1.92 × 10□ J) and last training sessions (5.05 × 10□ J), post-exercise MVC_ISO_ torque loss remained unchanged (Figure 3B). This observation is consistent with finding of a previous study, who demonstrated reduced strength loss during repeated eccentric cycling sessions for a similar workload (35). However, inter-individual variability in strength loss after the last training session was observed, with some participants exhibiting pronounced fatigue responses. This variability may be explained by the standardized progression of training workload, which did not account for individual differences in fatigue tolerance and may have led to excessive loading in some participants. Together, these results suggest that eccentric cycling induces robust adaptations that protect against exercise-induced fatigability, even as workload increases, although individualization of training load remains necessary to account for inter-individual differences in fatigue tolerance.

The high levels of mechanical strain absorbed during eccentric cycling training, enabled by the greater force production capacity during eccentric contraction (4, 13), provide a potent stimulus for improving maximal torque production on locomotor muscles. Muscle force is determined by the neural and muscular properties of motor units and the passive characteristics of the connective tissues involved in force transmission from muscle fibers to skeletal structure. Accordingly, the present study showed that MVC_ISO_, MVC_ECC_, and MVC_CONC_ torque produced at 180°.s^-1^, as well as jump height, increased after training with no difference between cadences (figure 2 and table 3). In contrast, MVC_CONC_ torque produced at 60°.s^-1^ increased only in the group trained at 60 rpm. This finding partly contrasts with previous reviews suggesting broad transferability of strength gains from eccentric cycling across contraction modes (4), which did not account for testing velocity. Indeed, earlier work has demonstrated velocity-specific adaptations following strength training (37). Those functional strength adaptations were mediated by neural (increase in voluntary activation level; table 4) and peripheral (increase in evoked torque and hypertrophy of VL and RF muscles; table 5) functions. While eccentric training has been proposed to induce specific structural remodeling through the addition of sarcomeres in series (17), the present study did not detect architectural changes beyond increased muscle thickness. Indeed, it was suggested that skeletal muscle responds to mechanical lengthening by adding sarcomeres in series resulting in a specific hypertrophic pattern to eccentric training. This increase in muscle material in series is supposed to impact the maximum velocity of shortening of muscle fibers (18), but the present training period of eccentric cycling did not assess muscle performance during high velocity contractions. As peripheral adaptation appears after 4-5 weeks of training, it was possible that our 6-week training period in a specific eccentric functional pattern did not lead to sufficient adaptations compared to longer training (5, 15, 36). Moreover, measurement limitations of 2D ultrasonography, particularly at a single site, may lack sensitivity to detect modest or region-specific adaptations (16).

### Practical applications

A 6-week eccentric cycling training program effectively improved functional capacity, but no substantial cadence effect was observed when comparing training at 30 and 60 rpm. Eccentric cycling specifically enhanced efficiency during eccentric, but not concentric, submaximal pedaling. Importantly, the progressive increase in workload did not exacerbate DOMS or post-exercise torque loss, supporting the good tolerability of this training modality. Although internal load did not differ between cadences, individualized exercise prescription remains essential in rehabilitation settings, with careful monitoring of tolerance and cardiovascular responses. Based on the present findings, lower cadences may be particularly suitable in rehabilitation contexts, as they appear well tolerated and safe for patients (9, 33).

## Data Availability

All data produced in the present study are available upon reasonable request to the authors.

## Author Contribution

All authors designed the project and contributed to the development of the methodology and the revision of the article. AM contributed to the methodology, data analyses, and writing of the article and conducted the experiments and processed the data. All authors read and approved the manuscript.

## list of abbreviations

1^st^ and 18^th^: the first and the last training session of eccentric cycling
CE: cycling efficiency
CMJ: countermovement jump
DOMS: Delayed onset muscle soreness
Dt10: double stimulations applied at 10Hz
Dt100: double stimulations applied at 100Hz
ECC: Eccentric contraction
ECC_30_: Eccentric cycling performed at 30 rpm
ECC_60_: Eccentric cycling performed at 60 rpm
EMG RMSMAX/MMAX: RMS EMG during MVC was measured during 500 ms around the peak torque and divided by MMAX area
RF: rectus femoris
Rf: respiratory frequency
HR: heart rate
HRR: half-relaxation rate
INT: intermediate test
MMAX: maximal M-wave
MVC_CONC_: isokinetic maximal voluntary concentric knee extensor contractions at velocity of 60°.s^-1^
MVC_ECC_: isokinetic maximal voluntary eccentric knee extensor contractions at velocity of 60°.s^-1^
MVC_ISO_: maximal voluntary isometric knee extensor contractions
O2 equiv: energetic equivalent associated to respiratory ratio
PO: power output
POST: post test
PPO: peak power output
PRE: Pre test
PE: Perception of effort
Rpm: revolution per minute
RTD: rate of torque development
SJ: squat jump
TT: Thorstensson test
Tw: single stimulation
VAL: Voluntary activation level
VE: minute ventilation
VL: vastus lateralis
VO2: oxygen consumption
VO2_PEAK_: peak oxygen consumption

---

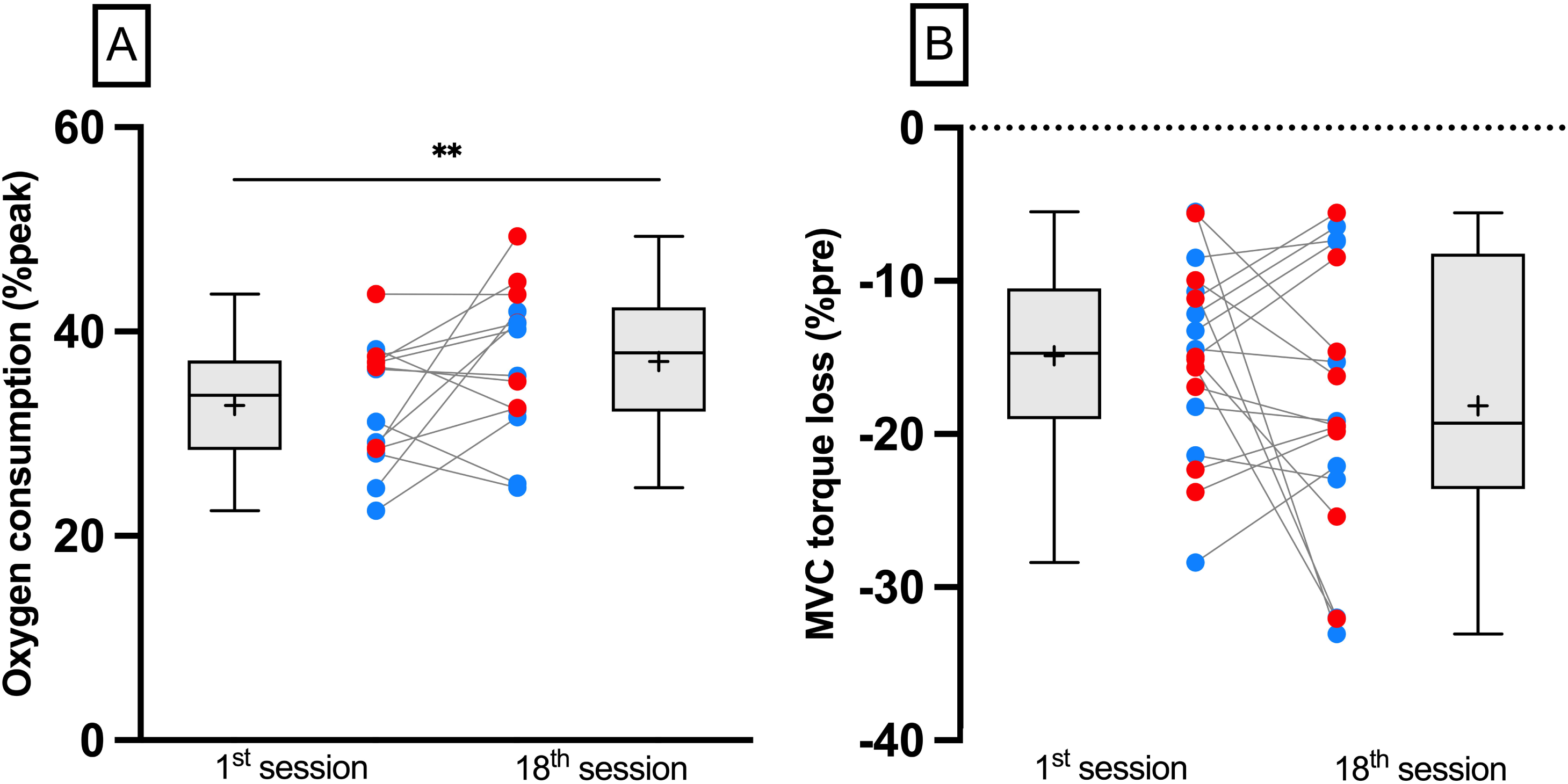

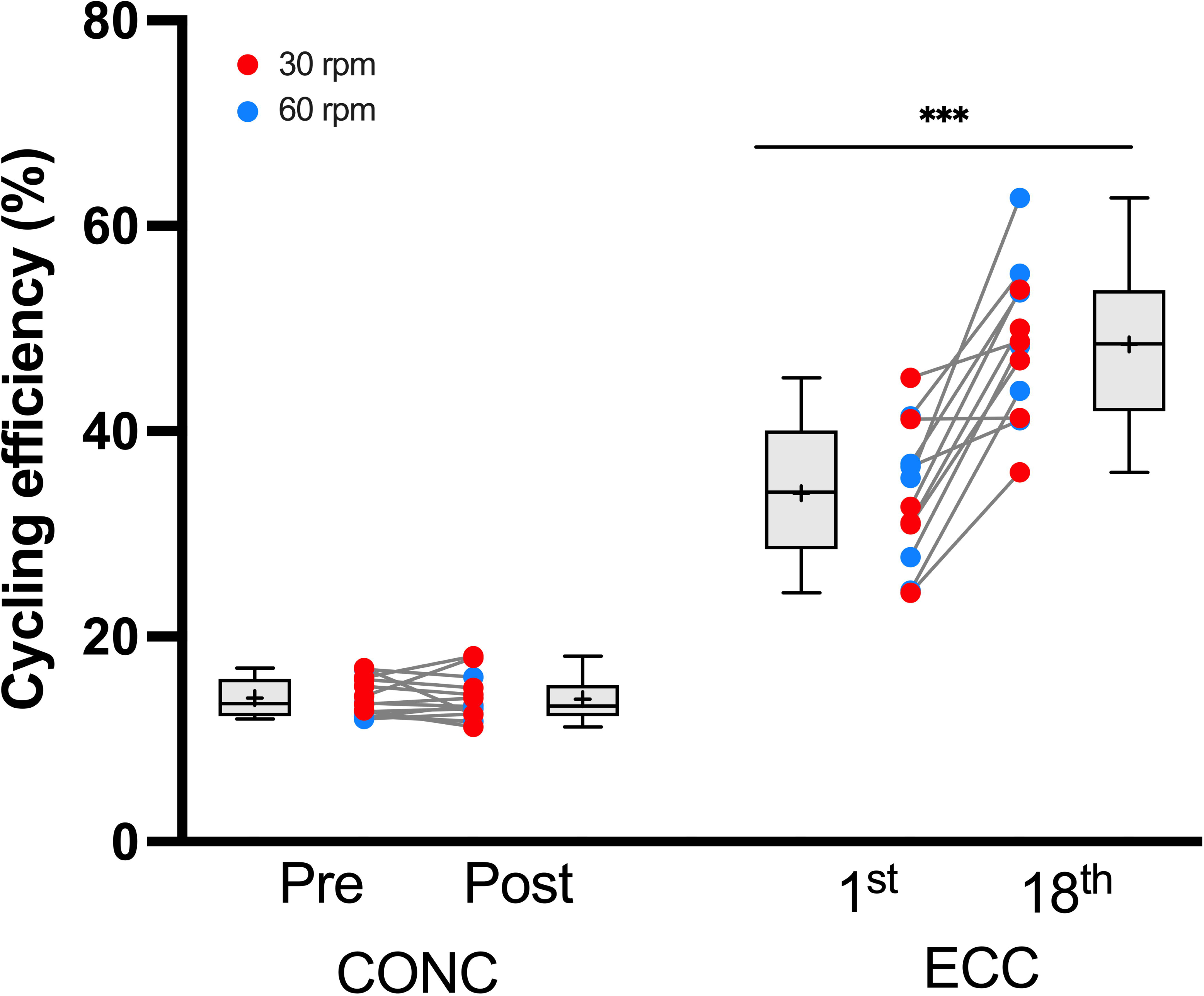

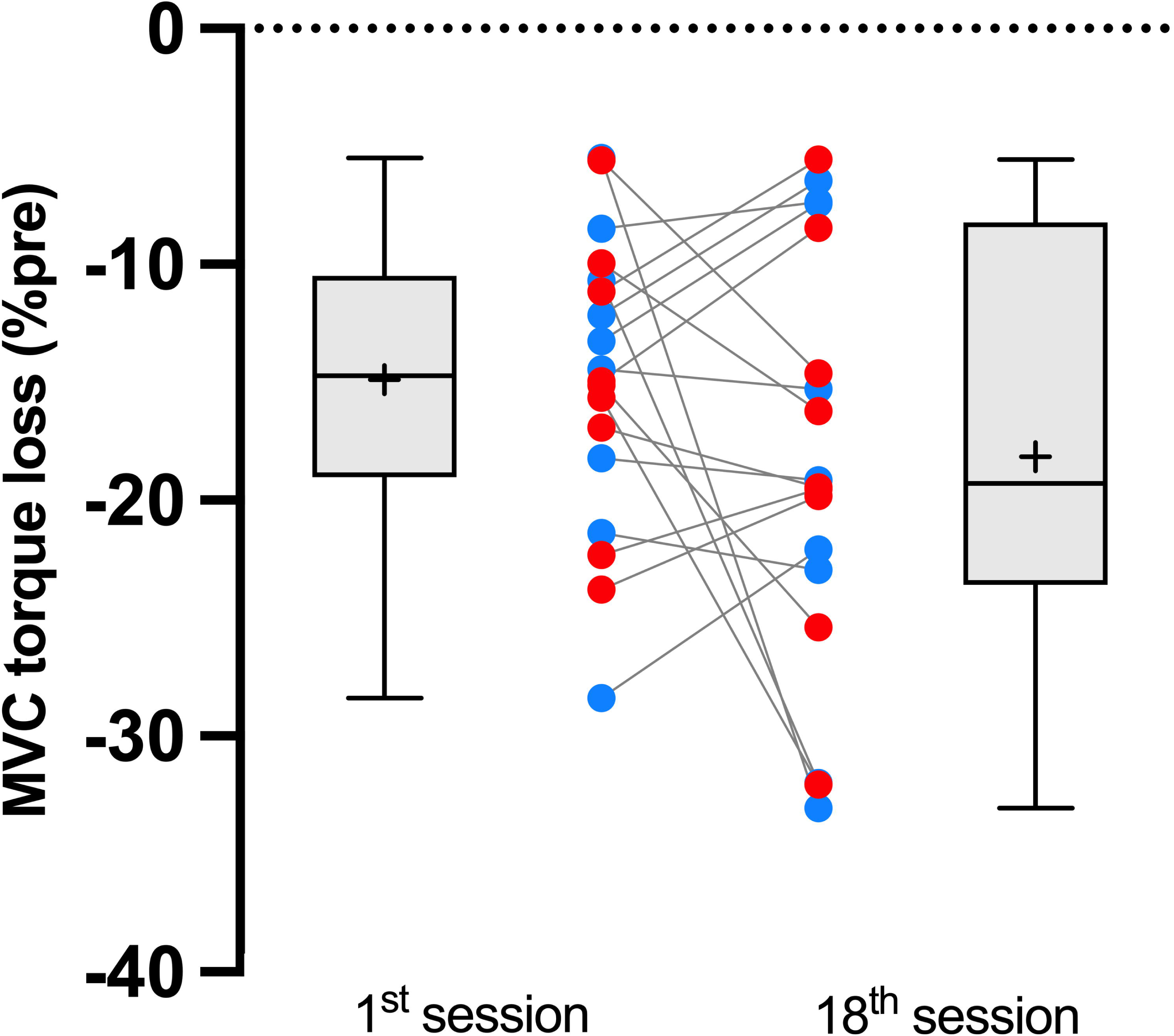

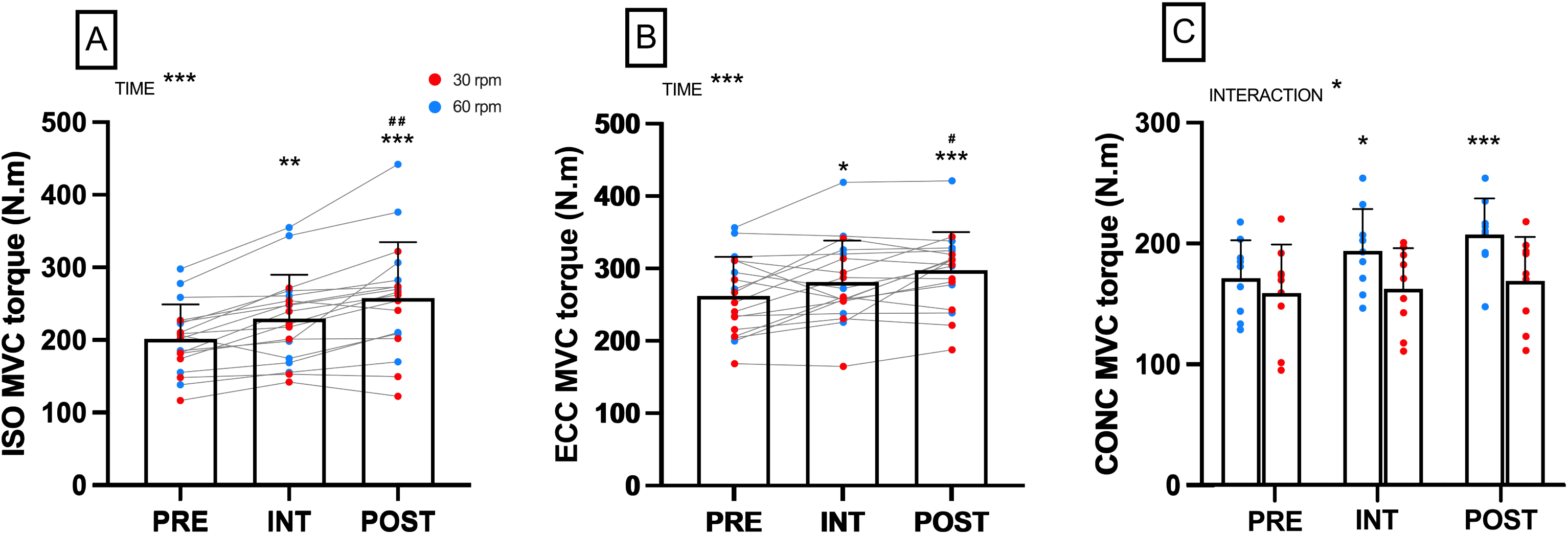

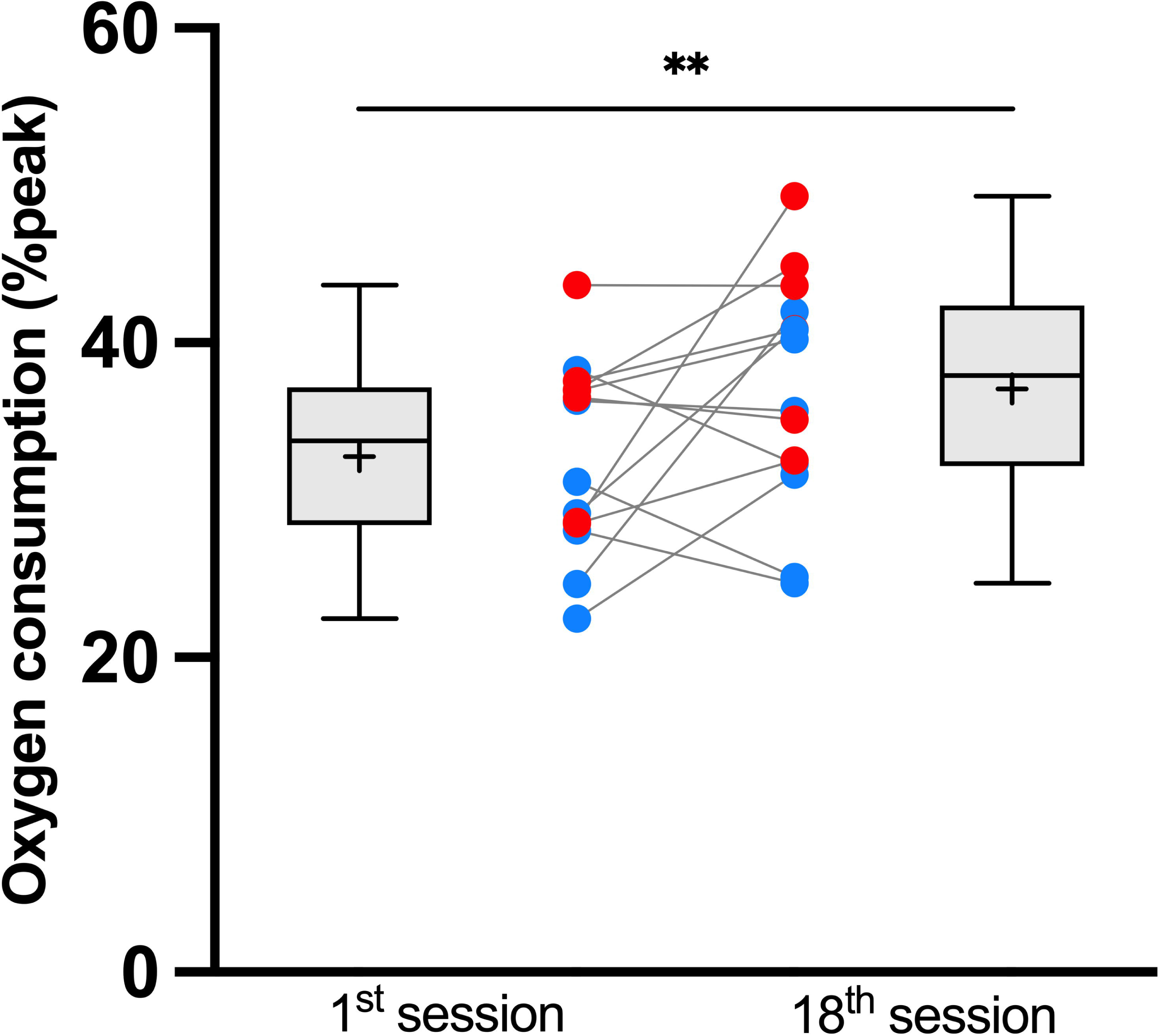

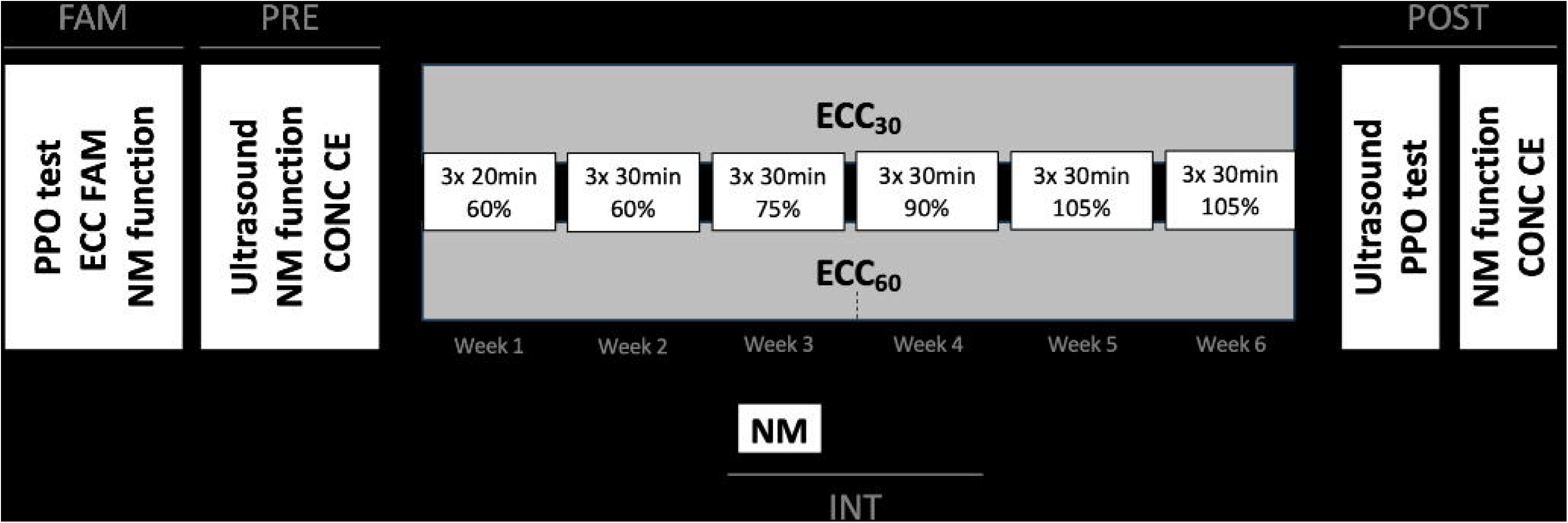

